# Vestibular contribution to path integration deficits in ‘at-genetic-risk’ for Alzheimer’s disease

**DOI:** 10.1101/2022.04.29.22274496

**Authors:** Gillian Coughlan, William Plumb, Peter Zhukovsky, Min Hane Aung, Michael Hornberger

## Abstract

Path integration changes may precede a clinical presentation of Alzheimer’s disease by several years. Studies to date have focused on how grid cell changes affect path integration in preclinical AD. However, vestibular input is also critical for intact path integration. Here, we developed a naturalistic vestibular task that requires individuals to manually point an iPad device in the direction of their starting point following rotational movement, without any visual cues. Vestibular features were derived from the sensor data using feature selection. Completing machine learning models illustrate that the vestibular features accurately classified *Apolipoprotein E* ε3ε4 carriers and ε3ε3 carrier controls (mean age 62.7 years), with 65% to 79% accuracy depending on task trial or algorithm. Our results demonstrate the cross-sectional role of the vestibular system in Alzheimer’s disease risk carriers and may explain individual phenotypic heterogeneity in path integration within this population

## Introduction

Path integration deficits manifest in early Alzheimer’s disease (AD) ^1^. Subtle changes in path integration are also present in adults at-genetic-risk of AD, decades before the expected onset of disease ^2–4^. Previous path integration studies have focused on visual cue-based tasks to investigate the behavioural and neural characteristic of the preclinical AD phenotype. However, vestibular input is critical for successful path integration, as it sends linear and angular information regarding self-motion from the inner ear to the brain via thalamocortical pathways ^5^ illustrated in Figure 1.

**Figure 1.**
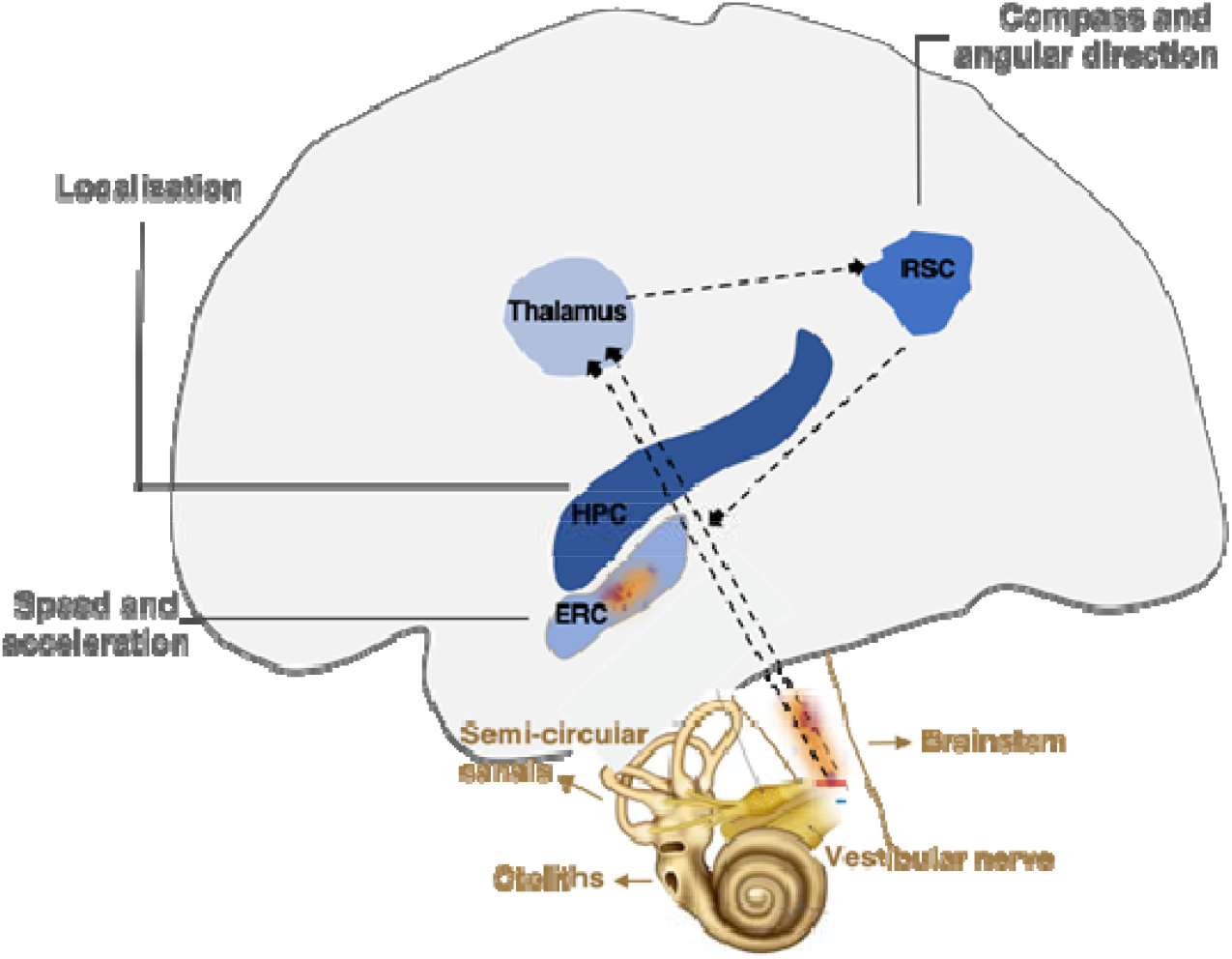
Hypothesised contributions of the vestibular system to path integration deficits in preclinical AD. The vestibular afferent nerve carries signals from hair cells in the semi-circular canals and otoliths of the inner ear to the brainstem. Projections from the brainstem carry vestibular signals via the anterior thalamocortical pathway to the thalamus and retrosplenial cortex, where directional information is processed by head direction cells. Information from the brainstem is also carried to the entorhinal cortex, where acceleration and speed information is processed. In Braak staging I and II, neurotoxic tau disposition appears in the brainstem and entorhinal cortex, potentially disrupting vestibular signalling to the neural system underlying path integration. ERC: entorhinal cortex; HPC: hippocampus; RSC: retrosplenial cortex. Black lines represent potential projections from one brain area to another.

We developed a real-world behavioural paradigm to measure vestibular function in ‘at-genetic-risk’ for Alzheimer’s disease. This task consists of three in-built vestibular sensors that record cardinal directions, acceleration (i.e., accelerometer) and rotational angle (i.e., gyroscope) during movement. This task was administered to apolipoprotein (*APOE*) ε4 allele carriers, of whom almost half develop Alzheimer’s disease by the average age of 76, compared to only 20% of non-carriers who convert to Alzheimer’s disease by the average age of 84 ^6^. Latent vestibular features were generated from the task data based on current theoretical knowledge and served as proxy measures of primary vestibular graviceptors, including angular velocity, which relies on intact function of the semi-circular canal of the inner ear and linear acceleration which relies on intact function of the otolith organ (see Table 1). Competing machine learning algorithms with five-fold cross-validation were used to test our hypothesizes that the vestibular features would play an important role for path integration performance in at-genetic-risk of AD.

**Table 1:** Vestibular Features

| Features (metric of measurement) | Feature description |
| --- | --- |
| *Path integration* |  |
| End Error (°) | This is the only feature not derived from motion and measures of basic PI performance. Scores are calculated by subtracting the final response position (by the participant) from the correct position like drop error traditionally used in navigation tests of PI (termed drop error in Kunz *et al.*, 2015a). |
| *Vestibular - semi-circular canal* |  |
| Total Angular Displacement (°) | Total angular displacement is the sum of all changes in compass points. For example, if a participant begins at 90 degrees and returns to zero, the perfect movement will have a total angular displacement of 90. If participants take a longer route in their motion response or turn the iPad backward and forward then the displacement will increase similar to end error measured during PI. |
| Tilt i.e. Largest Gyroscopic Value (°/s) | This feature calculates the largest (greatest magnitude +/-) gyroscopic value for each trial. Tilt orientation represents is a function of vestibular system that modulates PI performance (Angelaki *et al.*, 2009). This feature represents change of tilt orientation during movement response. Each gyroscope data point was taken every 0.003 seconds. |
| Quick vs longer Tilt i.e. Gyroscopic Rate of Change (°/s) | Average rate of change of tilt on the gyroscope data was calculated for each axis. For the rate of change, time intervals of 0.1, 0.5 and 1.0 seconds were used. For each time, intervals add all raw gyroscope data and divide it by the number of samples in the time interval. This decreases the amount of noise that might occur within the small refresh rates of the gyroscope by averaging. The reasoning for the different refresh rates is to evaluate if the participant is doing quick tilts or longer tilts. |
| *Vestibular - otolith organ* |  |
| Acceleration (m/s<sup>2</sup>) | The average acceleration feature represents the average of the accelerometer data during response movement, similar to time taken (i.e. duration) to complete PI during self-motion on virtual PI (similar to wayinfinfing duration; see Coughlan *et al.*, 2019). |
| Jerk (m/s<sup>3</sup>) | Average jerk is the time derivative of acceleration (de Winkel *et al.*, 2020). For each axis, a set amount of raw acceleration in a time (0.1, 0.5, 1.0) was summed, and the difference between each interval was calculated to generate the jerk value. |
| Hesitations | Average hesitation represented the accumulative number of stop and start movements in any one trial. This was calculated by using the moving window transformation of raw accelerometer data (peak detection function) to remove the noise in the raw data. |

The first feature represented basic path integration performance. The six vestibular features were dissociated based on the two the vestibular organs: the semi-circular canals that detects angular acceleration of the head and the otolith that detects linear acceleration. x-axis represents forward/backwards movement; y axis responses left/right movement; z axis represents up/down movement. PI: path integration.

## Materials and methods

### Participants and procedure

One hundred and fifty participants between 50 and 75 years of age were recruited to participate in a research study at the University of East Anglia, Norwich, UK. Written consent was obtained from all participants and ethical approval was obtained from Faculty of Medicine and Health Sciences Ethics Committee at the University of East Anglia, Reference FMH/2016/2017–11. Participants were a history of psychiatric or neurological disease, substance use disorder or motor control disorder were excluded. Participants receiving anti-depression or anti-anxiety medication at the time of screening were excluded. Saliva kits were sent to participants home and returned to the university on the same day the saliva sample was taken to determine *APOE* genotype status. Sensor data was collected on the iPad-based assessment tool (see *The Vestibular Rotation Task* for details), This data was only collected during the follow-up visit of the study, 18-months after the baseline assessments that are published. The final sample size of 53 included 32 ε3ε3 carriers and 21 ε3ε4 carriers at the cross-section each of whom completed the background cognitive testing and the vestibular task on the same day. We included a third genetic subgroup of homozygous *APOE*-ε4ε4 carriers in the supplementary materials, because they are very rare (*n* = 3).

### APOE genotyping

A saliva sample was collected for direct genotyping of APOE (with ε2 carriers removed). See (Coughlan et al., 2019) for further details

### Vestibular Rotation Task

The development of this vestibular task builds on the work of Mittelstaedt and colleagues (Figure 2) ^7^. Thus, to isolate the input of vestibular signals to performance, the task was administered in the complete absence of external visual or auditory cues. During the task, the participant was blindfolded and given earplugs on to ensure the test tapped into the vestibular system with no external stimuli. Participants sat in the rotating chair (feet not touching the floor) and held an iPad flat in their hands. Participants were asked to note set reference point in front of them, which was recorded on the iPad. During the trials, the examiner rotated the participant in the chair (see Table 2 for trial details) and remained behind them. Three seconds following the turn completion, participants were asked to point the iPad as accurately as possible in the direction of the reference point, while still wearing their blindfold and earplugs. The distance between the reference point and participant response (i.e. the end error) was recorded. This metric was used as a proxy measure of path integration, like previous studies^2,4^. During the pointing back movement response, the iPad sensors recorded accelerometer, gyroscopic, and compass information along the x-axis (forward/backwards motion) y axis (left/right motion) and z axis (up/down motion) (AppleInc 2021). See supplementary for an extend description of the task.

**Figure 2.**
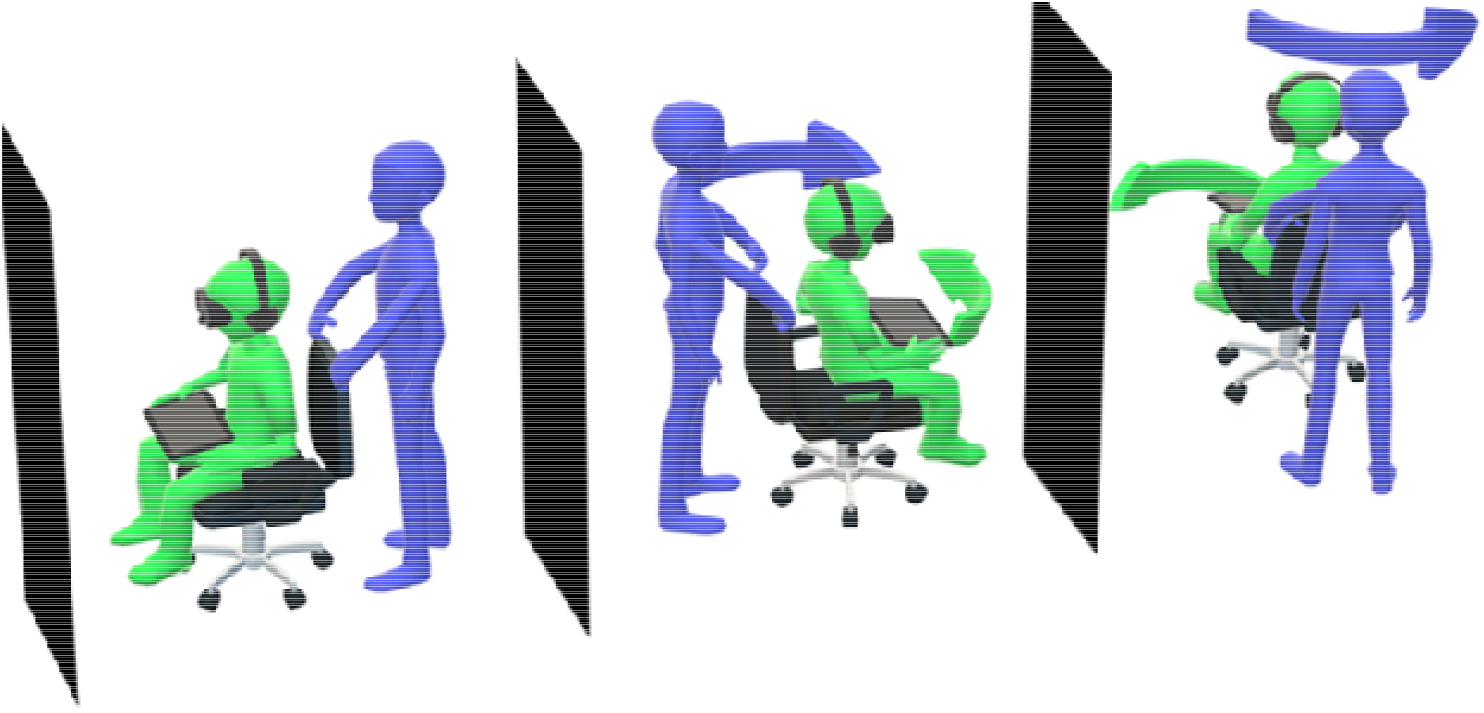
Vestibular Rotation Task. Participants were asked to raise their legs (i.e. not touching the floor) and were rotated by the tester (purple arrow). Three seconds following the turn completion, participants were asked to point the iPad as accurately as possible in the direction of the starting point, while still wearing the blindfold and earplugs (green arrow). As participants point back, the iPad recorded vestibular data: accelerometer (acceleration), gyroscope (rotation) and compass (direction). The experimenter then tapped the iPad to record the participants end location response which was later used to calculate end error. The tester always remained behind the participant to ensure that they did not become a location cue. To avoid direct feedback following each trial, the tester moved the participant back to the reference point before they remove the goggles (blindfold) and earplugs. Cardinal directions along which motion is recorded. Motion responses are automatically recorded along the x-axis (forward/backwards motion), y axis (left/right motion) and z axis (up/down motion) on the iPad (AppleInc 2021).

**Table 2:** The Idiothetic Rotation Task Rotations

| Trial | Degree of movement |
| --- | --- |
| 1 | 90 |
| 2 | 90 ⇒ -210 |
| 3 | 120 |
| 4 | 300 |
| 5 | 145 ⇒ -230 |
| 6 | 230 ⇒ -130 |
| 7 | -225 ⇒ 290⇒60 |
| 8 | -290 ⇒ 265 ⇒-70 |
| 9 | -200 ⇒ 310 ⇒-190 |

Degree of movement represents the rotations in each trial. Positive and negative values represent clockwise and anticlockwise rotations, respectively. Multiple rotations are separated by ⇒ symbol (i.e., multiple rotations before the participant points back to the starting point).

### Vestibular Features

The sensor data that was derived from accelerometer, gyroscope, and compass underwent pre-processing steps that are detailed in the supplementary. Following the data preparation phase, feature selection was used to derived meaning vestibular features from the task sensor data ^8^. This process was guided by theoretical knowledge on path integration, the vestibular system and cognitive impairment ^3,5,9–11^. Vestibular features are listed in Table 1. A data pre-processing procedure that preceded feature selection is detailed in the supplementary materials.

### Statistical analysis

Linear models were first applied to understand the extent to which univariate statistics would yield any detectable differences in vestibular features between genetic groups. Consistent with previous evidence that path integration deficits could be detected using a univariate approach, we expected differences on end error between the genetic groups, adjusting for age, sex and population. We did not expect to see differences between the genetic groups on the vestibular features derived from the sensor data.

### Machine learning algorithms

To determine whether vestibular features could distinguish between the genetic groups, classification models were created for each task trial. Each trial involves a different rotational movement, and it is not possible to combine all the trials. Classification accuracy of three competing machine learning algorithms [including Random Forest (RF)^12^, Support Vector Machines (SVM) ^13^and Multi-Layer Perceptron (MLP) ^14^] were computed via a standard class metric, the F1 score. The F1 score is a measure of the model’s performance that considers precision (true positives/true positives + false positives) and recall (true positives/true positives + false negatives):

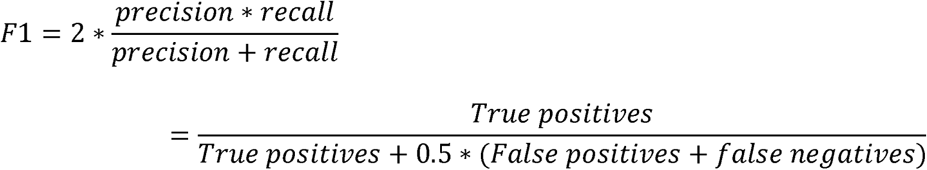

Thus, F1 score determined the classification performance for *APOE* status (i.e., ε3ε3 vs ε3ε4). A random classification F1 score is equal to 0.57, with a score of above 0.5 suggesting improved efficacy/sensitivity. Accuracy scores were also calculated as a secondary metric and represents the percentage of correctly classified ε3ε3 and ε3ε4 participants. Two sets of analyses were performed: in the first set of models, all features were included (blue line in Figure 4A). In the second set of models, end error was excluded to assess the independent precision of vestibular function to the prediction of genetic risk (red line in Figure 4A). An overview of the machine learning algorithms is provided in Supplementary.

### Cross-validation

A control validation resampling procedure evaluated the machine learning models ^15^. This method is typically used on a limited data sample. Five-fold validation was used using four groups as training data and one for testing. Five F1 scores for each algorithm and for each trial were computed and were averaged for validity. Each of five groups are created using stratified folds to preserve the samples in both *APOE* groups.

### Data availability

The raw and pre-processed data, as well as the code for the machine learning algorithms, are available at: https://github.com/WillPlumb/Vestibular-PreAD

Demographic participant data are available on request

## Results

### Demographic characteristic and univariate statistics

Demographic and neuropsychology characteristics were not significantly different between genetic groups (Table 3). Secondary characteristics are presented in Table S1. Path integration significantly differed between groups (*P*=0.02) on Trial 1, with ε3ε4 carriers showing a greater degree of response error compared to ε3ε3 carriers (Figure 3A). This effect was not significant on trials 2-9, when novelty induced by initial exposure to the task attenuated. An overview of the t-statistics for each feature on each trial is shown in (Figure 3B), showing that linear models do not detect differences in vestibular function as a function of genetic group.

**Figure 3.**
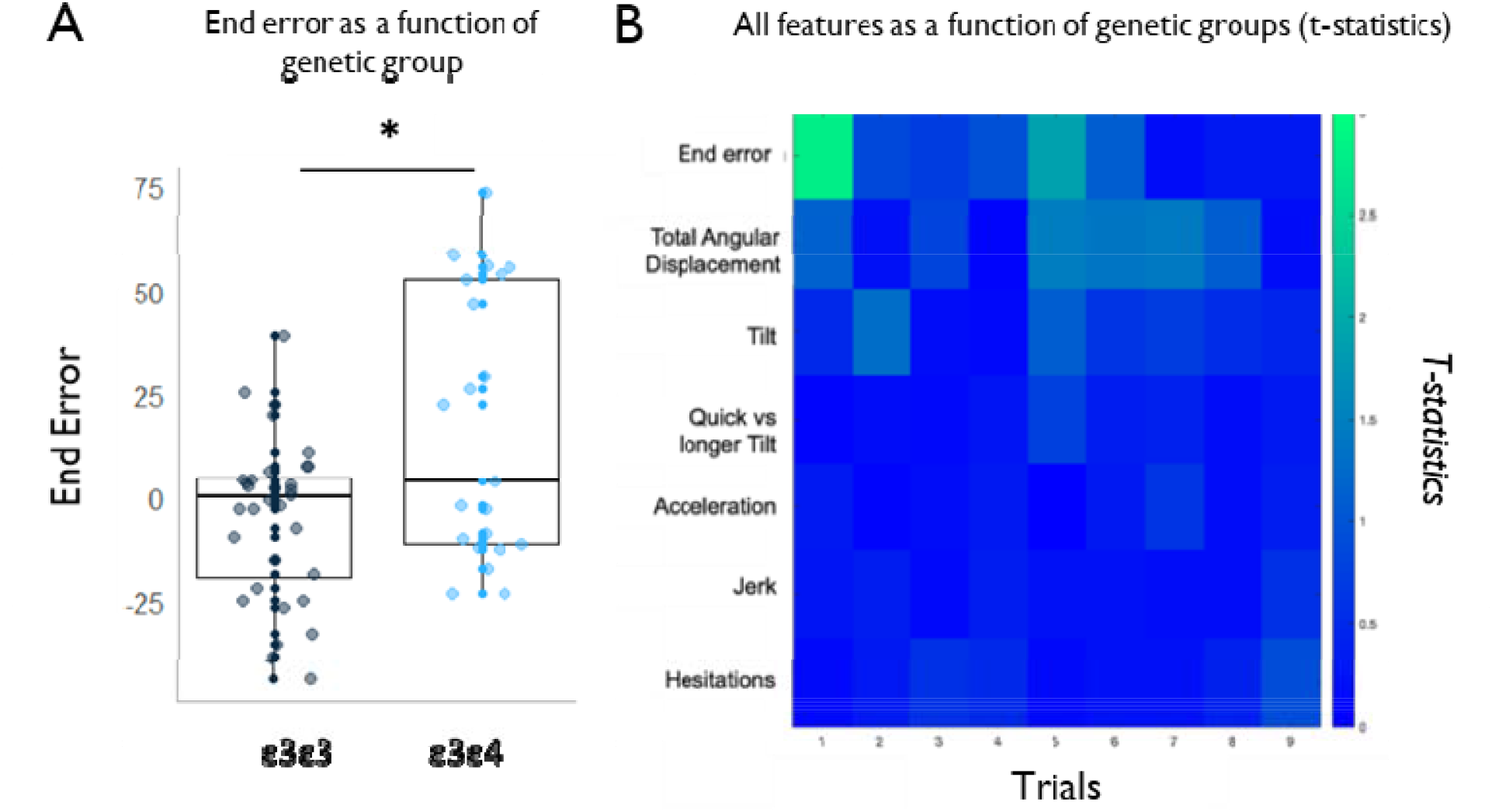
Univariate statistics and machine learning results. **A** *APOE* ε3ε4 carriers had a higher rate of end error compared to the ε3ε3 carriers in Trial 1. **B** No statistical differences in the mean values for each vestibular feature, adjusting for age sex and occupation. x, y and z axis are collapsed for the visualisation.

**Table 3:**
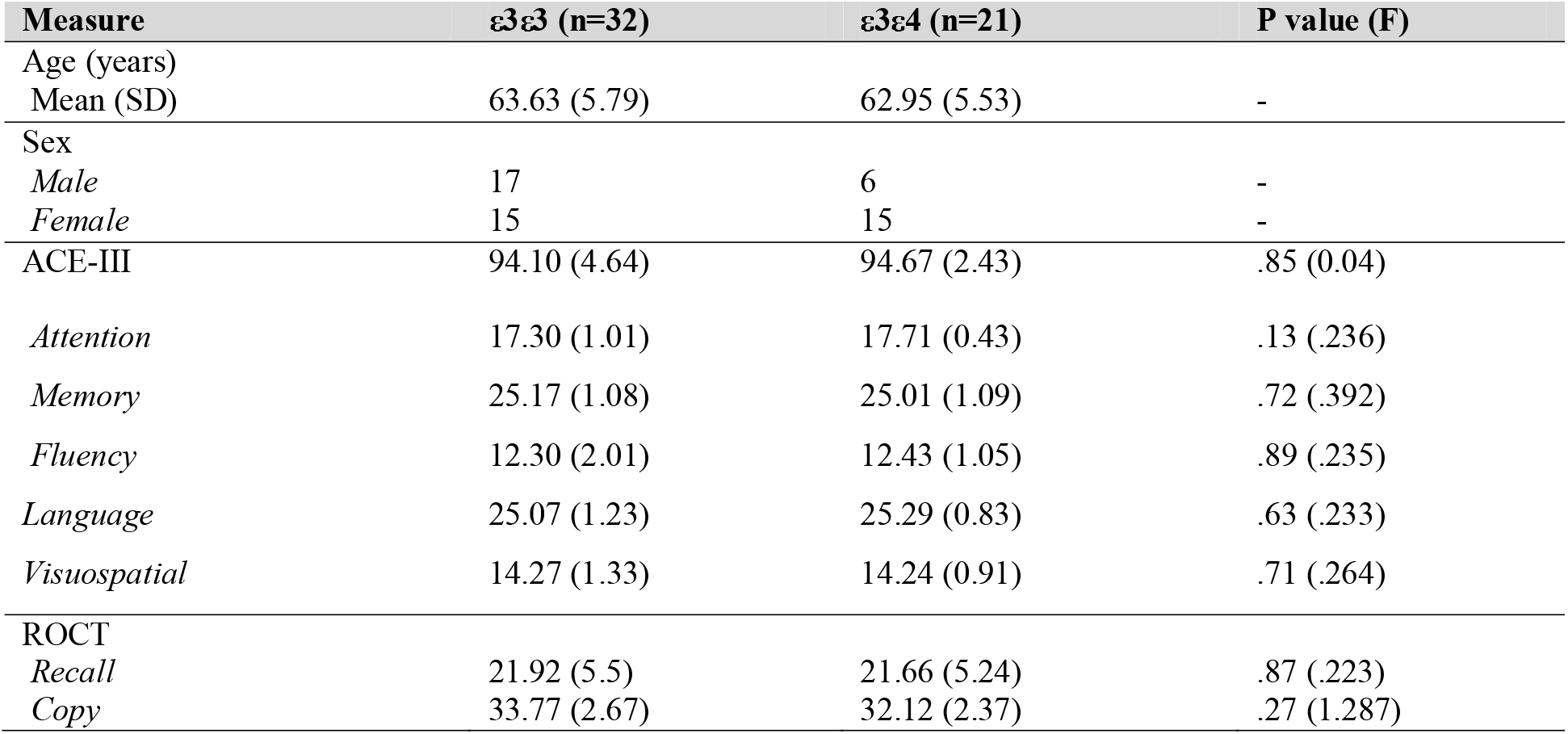
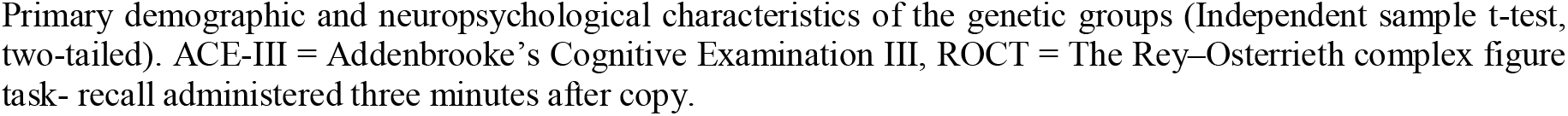
Primary demographic and neuropsychology characteristics of the sample

| Measure | ε3ε3 (n=32) | ε3ε4 (n=21) | P value (F) |
| --- | --- | --- | --- |
| Age (years) |  |  |  |
| Mean (SD) | 63.63 (5.79) | 62.95 (5.53) | - |
| Sex |  |  |  |
| Male | 17 | 6 | - |
| Female | 15 | 15 | - |
| ACE-III | 94.10 (4.64) | 94.67 (2.43) | .85 (0.04) |
| Attention | 17.30 (1.01) | 17.71 (0.43) | .13 (.236) |
| Memory | 25.17 (1.08) | 25.01 (1.09) | .72 (.392) |
| Fluency | 12.30 (2.01) | 12.43 (1.05) | .89 (.235) |
| Language | 25.07 (1.23) | 25.29 (0.83) | .63 (.233) |
| Visuospatial | 14.27 (1.33) | 14.24 (0.91) | .71 (.264) |
| ROCT |  |  |  |
| Recall | 21.92 (5.5) | 21.66 (5.24) | .87 (.223) |
| Copy | 33.77 (2.67) | 32.12 (2.37) | .27 (1.287) |

Primary demographic and neuropsychological characteristics of the genetic groups (Independent sample t-test, two-tailed). ACE-III = Addenbrooke's Cognitive Examination III, ROCT = The Rey–Osterrieth complex figure task- recall administered three minutes after copy.

### Vestibular features distinguish ε4 carriers from non-carrier controls

Next, we sought to examine if the vestibular features distinguished the genetic groups by applying a machine learning approach. The three choosen algorithms examined the classification performance of the vestibular features for APOE genotype status. The vestibular features are listed in Table 1. Six out of the nine trials achieved a cross-fold average F1 score of above 0.6 using one or more of the algorithms, suggesting stable differences in vestibular function among *APOE* ε4 carriers and non-carriers. Accuracy percentages ranged from 65% to 79% depending on task trial and algorithm. Excluding age, sex and occupation data produced similar F1 scores, suggesting that vestibular function classified *APOE* status irrespective of demographic variation (see Table S2). Please refer to Table S3 for results including a small number of highest-genetic-risk ε4ε4 carriers who were included in the *APOE* ε4 carrier group, subsequently producing greater F1 scores (reaching 0.77) across all trials. We then examined the influence of the vestibular function when the path integration proxy end error was excluded. Excluding this feature led to a best prediction accuracy of 0.75, suggesting that vestibular function alone maintains good classification performance for *APOE* status. Accuracy ranged from 65%-75%. The highest performing algorithm for each trial including and excluding path integration is presented in Figure 4A. F1 scores and accuracy scores across all trials and best performing algorithm are presented in Table 4.

**Figure 4.**
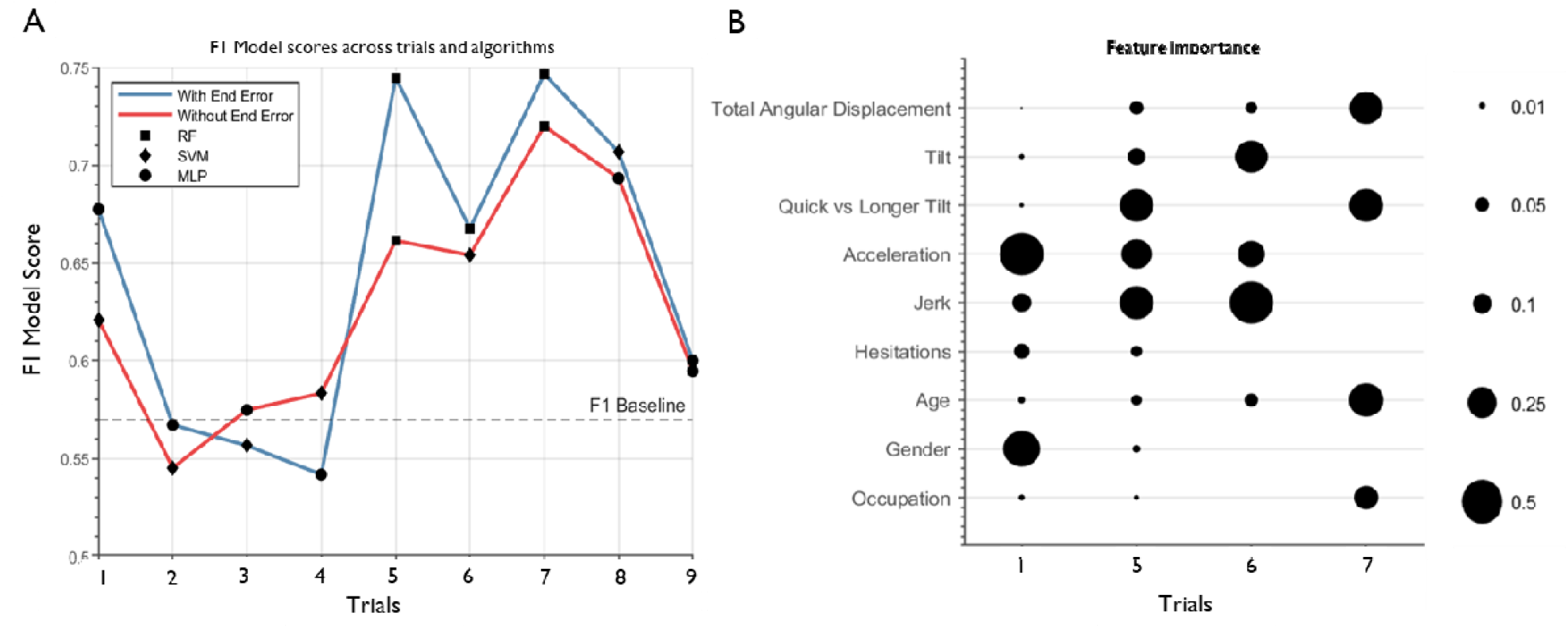
Feature importance. **A** F1 scores for the best performing algorithm are shown. A random predictor would score 0.57, with a score of above 0.57 representing better-than-chance APOE status classification performance. Blue line includes all features. Red line excludes the path integration feature, end error. **B** Importance scores are represented by the circle diameter and were derived for the best performing model on each of the trials shown. Scores vary between 0 and 1 depending on the proportion of influence the feature has for that trial. RF=Random Forest, SVM= Support Vector Machine, MLP=Multi-Layer Perception

**Table 4:** **A** All features including demographics age, sex and occupation and all engineered features. **B** Precision when the only ‘non-motion-based’ path integration feature; end error was excluded. F1 scores reflect the best performing models and algorithm on each trial. A random predictor would score 0.5 or lower, with a score of above 0.5 representing task prediction efficacy. RA=Random Forest, SVM= Support Vector Machine, MLP=Multi-Layer Perceptron.

| Trial | **A. All features** |  | **B. Excluding end error** |  |
| --- | --- | --- | --- | --- |
|  | Best F1 Score | Algorithm | Best F1 Score | Algorithm |
| 1 | 0.677 | MLP | 0.621 | SVM |
| 2 | 0.567 | MLP | 0.545 | SVM |
| 3 | 0.557 | SVM | 0.575 | MLP |
| 4 | 0.542 | MLP | 0.583 | SVM |
| 5 | 0.745 | RF | 0.661 | RF |
| 6 | 0.669 | RF | 0.654 | SVM |
| 7 | 0.747 | RF | 0.720 | RF |
| 8 | 0.707 | SVM | 0.693 | MLP |
| 9 | 0.600 | MLP | 0.595 | MLP |

### Identifying the most influential vestibular features

The identified features tapped into both vestibular pathways and varied across task trials, suggesting that vestibular pathways influencing the path integration brain network may be similarly influenced by the *APOE* ε4 variant (Figure 4B). Demographics are included as a means of comparison.

## Discussion

Our results show a vestibular function impairment in at-genetic-risk of AD. Vestibular function differentiated ε3ε4 carriers from ε3ε3 carriers, irrespective of demographic background. Machine learning algorithms achieved good performance (F1 and accuracy scores of up to 0.72 and 0.75, respectively) for classifying genetic groups based on vestibular function, while univariate statistics failed to identify vestibular differences between the *APOE* groups. We also replicate previous path integration deficits in *APOE* ε4 gene carriers when task novelty is high^3,4^. Our findings highlight the need for both more naturalistic cognitive tasks and a broader computational perspective to understand the preclinical AD phenotype.

### Dysregulation of the vestibular system is associated with path integration deficits

Evidence from animal models has consistently demonstrated the importance of the vestibular system to path integration. Vestibular lesions impair spatial memory and the ability to rerun to a goal location following passive transport ^16^. This effect is exacerbated in darkness ^17^. Interestingly, rodents with vestibular lesions path integrate successfully with the aid of external visual cues ^18^. This has striking similarities to Bierbrauer and colleagues who found that a path integration deficit in *APOE* ε4 carriers emerges only if external visual cues are not available ^4^. These findings are also consistent with limited studies that show path integration impairment in vestibular deficient humans emerges when external cues are not available^19^, supporting the emerging theory that vestibular function plays a mechanistic role in path integration deficits previously observed in adults at-genetic-risk AD.

### Dysregulation of the vestibular system is associated with spatial cell dysfunction

Evidence from studies in animals and humans also suggests strong anatomical and functional interdependency between the vestibular system and the navigation system. The entorhinal cortex receives inputs from the vestibular system via the anterior thalamic nuclei and the retrosplenial cortex along the anterior thalamocortical pathway ^5,20–23^. Peripheral vestibular input impacts head direction during self-motion, which relies on head direction cells and degradation of head direction cell activity can be caused by loss of vestibular information from the otolith organs and the semi-circular canals ^24,25^. Disruption of the head direction cell network impairs the grid cell signal in the entorhinal cortex and parasubiculum, which are crucial for path integration^26^. Moreover, hippocampal place cells that code for specific environmental locations ^27^, are partially modulated by vestibular stimulation^28,29^. Disruption of the vestibular input to the hippocampus via bilateral labyrinthectomy eliminates place cells firing, interfering with self-movement signals^18^. Interestingly, altered task-related hippocampal activity has also been linked to path integration impairment in adult *APOE* ε4 carriers^2^. However, this hippocampal activity is increased and correlates with better spatial memory. Thus, vestibular projections may dysregulate the grid cell signal in at-genetic-risk of AD, although this remains to be tested. In sum, these data point to a mechanistic role of vestibular signalling leading spatial cell dysfunction at-genetic-risk AD.

### Clinical implications

Vestibular signals that influence path integration in preclinical AD may help pinpoint pre-disease pathological changes and in turn guide treatment. Identifying vestibular contributions to the cognitive phenotype of preclinical AD is important because vestibular dysfunction is often present with treatable hearing loss^30^, recently cited as a modifiable risk factor for AD^31^. Moreover, vestibular balance training such as Intensive Slackline-Training improves path integration and vestibular function and implanted vestibular prosthesis (that reproduces information normally provided by semi-circular canals) improves spatial orientation in monkeys with severe vestibular damage ^33^, suggesting adults with vestibular dysfunction (and path integration impairments), may respond to a vestibular implant and or vestibular intensive training. Moreover, because the vestibular system has extensive connections to AD vulnerable brain regions including the hippocampus, cingulate cortex and parietal lobe, vestibular stimulation may indeed improve cognitive performance related to the integrity of these brain regions, including disorientation and memory loss.

### Limitations, future research directions and conclusion

Vestibular contributions to the cognitive phenotype of preclinical AD warrant further investigation. Our study included an APOE genotyped sample that was a similar sample size to previous studies ^2,3^. Nonetheless, this sample is moderate in the context of machine learning approaches, even though we tried to remedy this with the inclusion of five-fold cross-validation of our classification models. Further research is needed to understand how degraded graviception contributes to deficits in self-motion perception and the role played by head direction and grid cell dysfunction in graviception. Our approach to investigating vestibular contributions should be considered a steppingstone towards more tailored treatment programs for preclinical AD, that can be combined with pharmacological, non-pharmacological sensory stimulation, gamma-induction, and dietary treatment strategies 34,35.

In conclusion, we introduce a novel task that measures human vestibular function and path integration. The application of machine learning to vestibular data reveals vestibular changes in at-genetic-risk adults, which should be a priority for basic scientific research on human vestibular function. Our findings should accelerate objective, high frequency and passive digital phenotyping of at-genetic-risk AD and help elucidate the mechanisms by which the human vestibular system contributes to cognitive impairment in preclinical AD. Future studies should examine firstly, whether unstable grid cell representations detectable in task-based fMRI are causally linked to vestibular dysfunction in at-genetic-risk AD and secondly whether vestibular changes precede, or succeed, the emergence of tauopathy in the brainstem and entorhinal cortex.

## Data Availability

The raw and preprocessed data as well as the code for the machine learning algorithms are available.

https://github.com/WillPlumb/Vestibular-PreAD

## Abbreviations

APOE: Apolipoprotein E
RF: Random Forest
SVM: Support Vector Machine
MLP: Multi-Layer Perception

## Acknowledgements

We thank Tom Percy for programming the sensor task and the many participants who gave their time to this research.

## Funding

Faculty of Medical at the University of East Anglia (GC, MH)

The Alzheimer Society of Canada (GC, 22-08)

The Alzheimer’s Research United Kingdom (MH)

Canadian Institutes of Health Research postdoctoral fellowship (PZ)

## Supplementary material

Supplementary material is available at *Brain* online

## Supplementary Materials for

### Materials and Method

#### Vestibular Rotation Task

The development of this vestibular task builds on the work of Mittelstaedt and colleagues ^1–4^. Vestibular signals interact with sensory signals such as visual optic flow or auditory perception during path integration^5^. Thus, to isolate the input of vestibular signals to performance, the task was administered in the complete absence of external visual or auditory cues. Decisions on rotation selection for each trial were made by considering level of task difficulty. During the task, the participant was blindfolded and given earplugs on to ensure the test tapped into the vestibular system with no external stimuli. Participants were invited to sit in the rotating chair and hold an iPad flat in their hands. The tester ensured that the participant had enough leg space and that their legs would not touch objects when completing the test, which could serve as an external localisation cue. Participants were asked to note a reference point in front of them, which was held constant throughout the trials. This point was then set as the reference point on the iPad for each trial. Note the position of the starting point varied from participant to participant due to the use of differing testing rooms. The tester rotated the participant in the chair (see Table 2 for rotation details in each trial). As inertial inputs are affected by velocity^2^, the participant was rotated by the tester aiming to ensure a constant and consistent speed across the trials. Three seconds following the turn completion, participants were asked to point the iPad as accurately as possible in the direction of the starting point, while still wearing the blindfold and earplugs. During the pointing back, the iPad records data from the difference sensors: accelerometer, gyroscope, and the compass. The tester always remained behind the participant to ensure that they did not become a location cue. The iPad task automatically recorded the responses when the tester tapped on the iPad screen immediately following the participants response. iPad responses are automatically recorded along the x-axis (forward/backwards motion) y axis (left/right motion) and z axis (up/down motion) (AppleInc 2021). The vestibular rotation task is presented in Figure 2. The vestibular information provided in the ascending pathways to the limbic system (including medial temporal lobe) are required for accurate internal representation of the relationship between self-location, before, after, and during movement ^6,7^. If vestibular signals are affected in at-genetic-risk AD, this internal representation is negatively altered and vestibular performance will be altered.

#### Pre-processing preceding feature extraction

Python (version 3.2) and R studio (version 1.4) were used for pre-processing and statistical analysis. A signal pre-processing stage was first applied to the raw accelerometer, gyroscope and compass sensor data in order to mitigate signal noise and instrumental artefacts. This in turn allowed us produce signals that represented accurate response movements, which were given by the participant at the end of each trial. Noise removal involved using moving window averaging filters to smooth by averaging over 0.1 seconds. We used a set of cleaning rules (also known as bespoke heuristics) to correct for sensor errors such as unnatural movements. For each participant, a different reference/start point was set on the iPad prior to the task. To enable analysis between each participant the starting orientation was set to have the same compass value. Subtracting each participants’ reference start point from their current location produced the start angle. If instantaneous changes in the raw signals occurred due to hardware-based errors in the raw compass data, sensor errors, or measurement errors, the implementation applied simple addition or subtraction at each change point. This removed large value changes caused by the hardware set at changes of over 80 degrees between sequential data points. This threshold was chosen as it near impossible that a participant would turn the iPad 80 degrees within 0.1 seconds and thus such changes must be a result of sensor artefacts. If the participants held the iPad reversed hence the movement was inverted, the values were mirrored so all movements were in the same direction. These steps ensured that all the heading data was comparable between participants and thus we calculated the orientation difference between the start points and current compass positions, which is comparable between the participants. A raw and processed data example is represented in Fig. S2. Following data processing, the participant numbers for each group (ε3ε3, ε3ε4) on each trial include: Trial 1 (32,21), Trial 2 (32,21) Trial 3 (31,21) Trial 4 (32,21) Trial 5 (19,16) Trial 6 (32,21) Trial 7 (11,10) Trial 8 (11,10) Trial 9 (31,21).

#### Feature collinearity

Following feature selection ^8^, Spearman’s correlations were used to assess multicollinearity between the vestibular features generated in Table 1 (see Fig. S1 for a heat map representation the correlation between features [x, y, and z axis of the iPad summed] on trial one). For the Spearman’s rank correlations for each pair of features, a correlation threshold was set to reduce multicollinearity. The method was initiated by starting with “end error” and evaluating all correlation values against it. If the correlation with a further feature does not exceed the threshold, then it is added to a subset of new subset of features with “end error”. Using the next feature in the subset, we check each pair of features and remove one that correlates over the threshold. This continued until all features pairs in the subset were evaluated (i.e. until all pairs of features in the subset are less correlated than the threshold value). We test this with a set of threshold values {spearman’s r: 0.05, 0.1, 0.2, 0.3, 0.5, 1.0} to gain multiple feature sets allowing us to evaluate the best feature set for the machine learning algorithms, with 0.05 being the most stringent threshold (no features in the prediction model) and 1.0 is the most relaxed threshold (all features in the model).

**Fig. S1.**
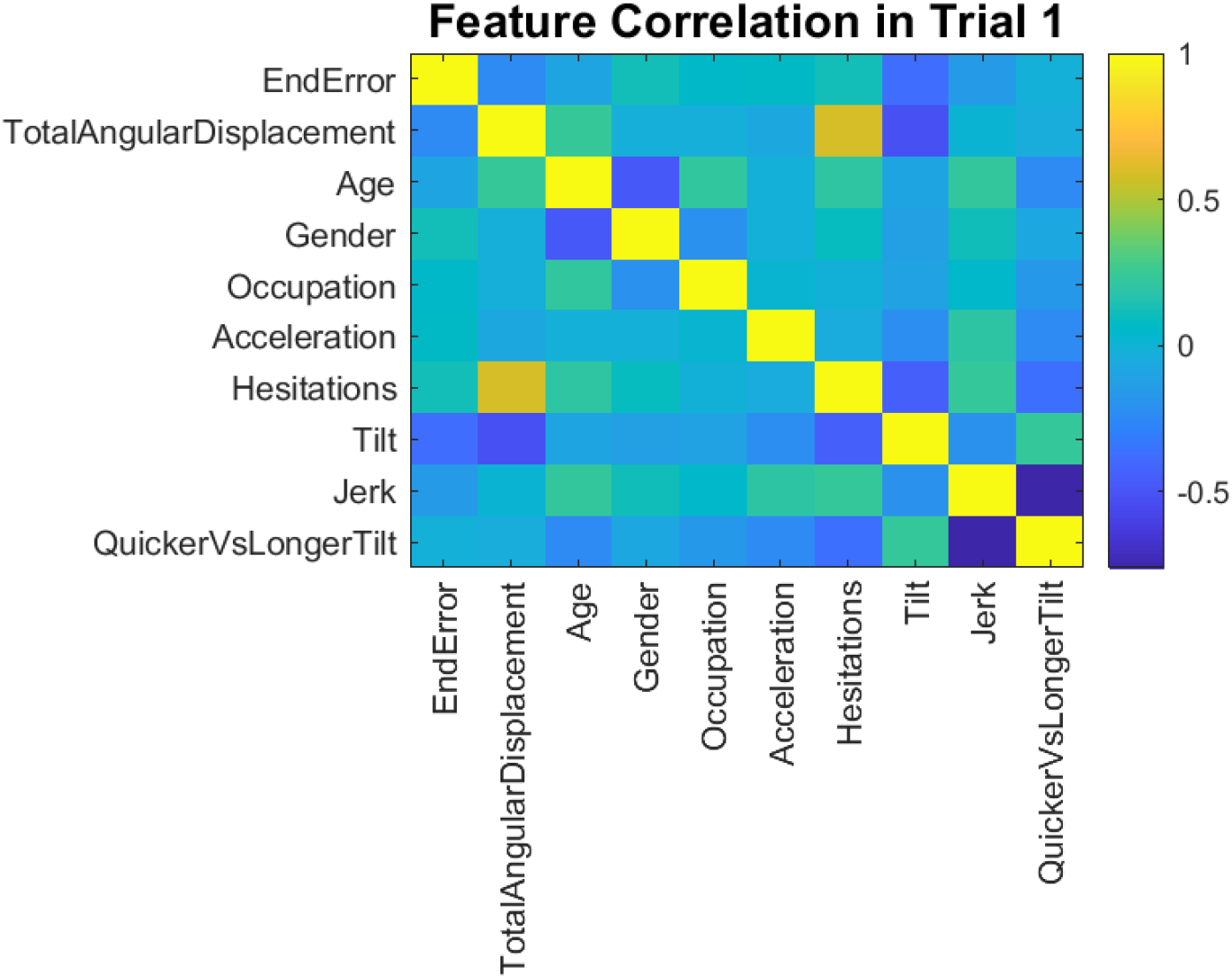
Correlation heat map for trial one using Spearman’s rank correlation. The correlation value was calculated for each pair of features to set a correlation threshold to remove any feature that correlates highly with other features. The correlation threshold was varied from 0.05, 0.1, 0.2, 0.3, 0.5, 1. The method is initiated by adding the first feature (end error) and then checking all correlation values against it. If any other feature breach this threshold, it is removed from the original list of features. The next feature in the original feature list is then selected until none remain.

**Fig. S2.**
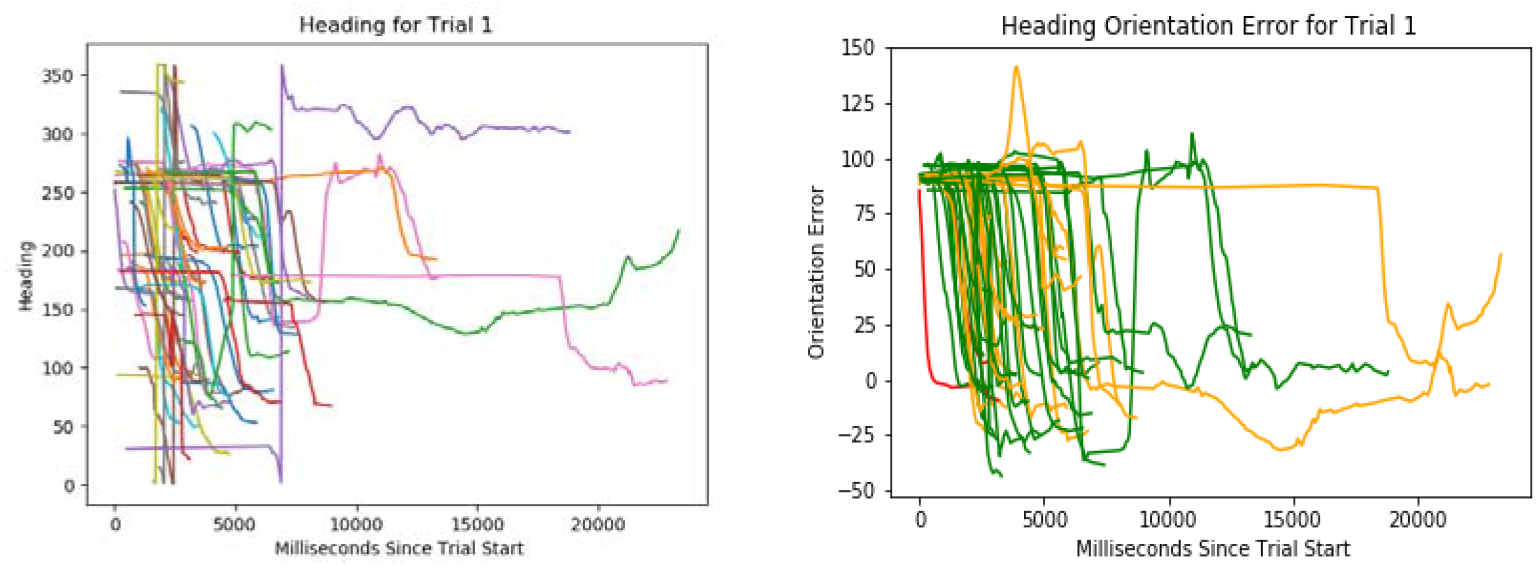
Example of signal pre-processing which allowed us to clean heading orientation from the raw compass signal (left) to the clean compass signal (right).

#### Machine learning algorithms

*The Random Forest Algorithm* (RF) comprises of decision trees and was chosen due to its applicability to large features sets via as pruning. A set of features is selected to train a decision tree based on the *APOE* genotype. The maximum number of features per tree and number of trees was set. Each individual tree selects features that best separate the *APOE* target, meaning less important features will not be used. The final model is used to benchmark any prediction results against, as it includes all features. Two variable parameters for the RF model were tested, the first being the number of decision trees used in the ensemble, where 100, 250, 500 and 1000 trees were tested. The second being the maximum number of features available to each tree. We applied the proportional factors 0.2, 0.5 and 0.7 which were multiplied by the total number of features.

*Support Vector Machine* (SVM) is a widely used type of classification model based on the optimization of an *N*-dimensional hyperplane, where *N* is the number of features. Linear and non-linear radial kernels were used. Values of the parameter *C* which controls the margin of the hyperplane was tested with the values (0.5, 1, 3, 5, 10 and 20).

*Multi-Layer Perceptron* (MLP) is a widely used feed forward neural network that can be applied to classification tasks. For the MLP, we tested three types of the activation functions of the neurons (identity function, logistic function and the hyperbolic function), the alpha constant, which is a parameter used to constrain the flexibility of the model to prevent overfitting {0.0001, 0.0005, 0.001, 0.002} and the number of nodes in the hidden layer {5, 10, 15, 20}.

**Fig S3.**
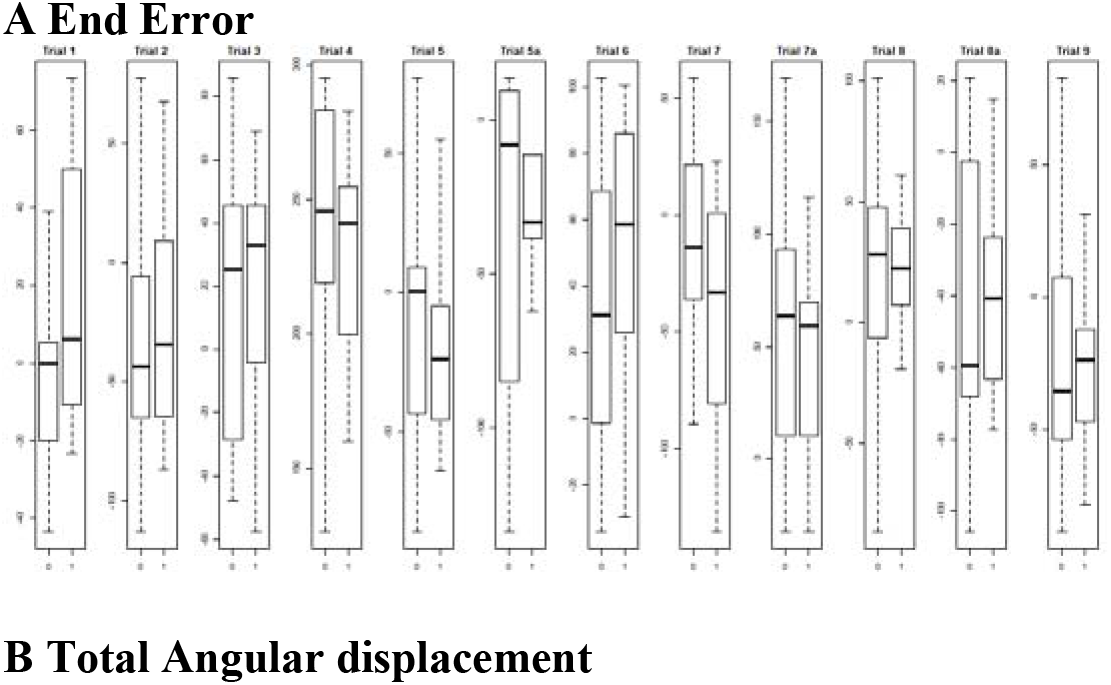
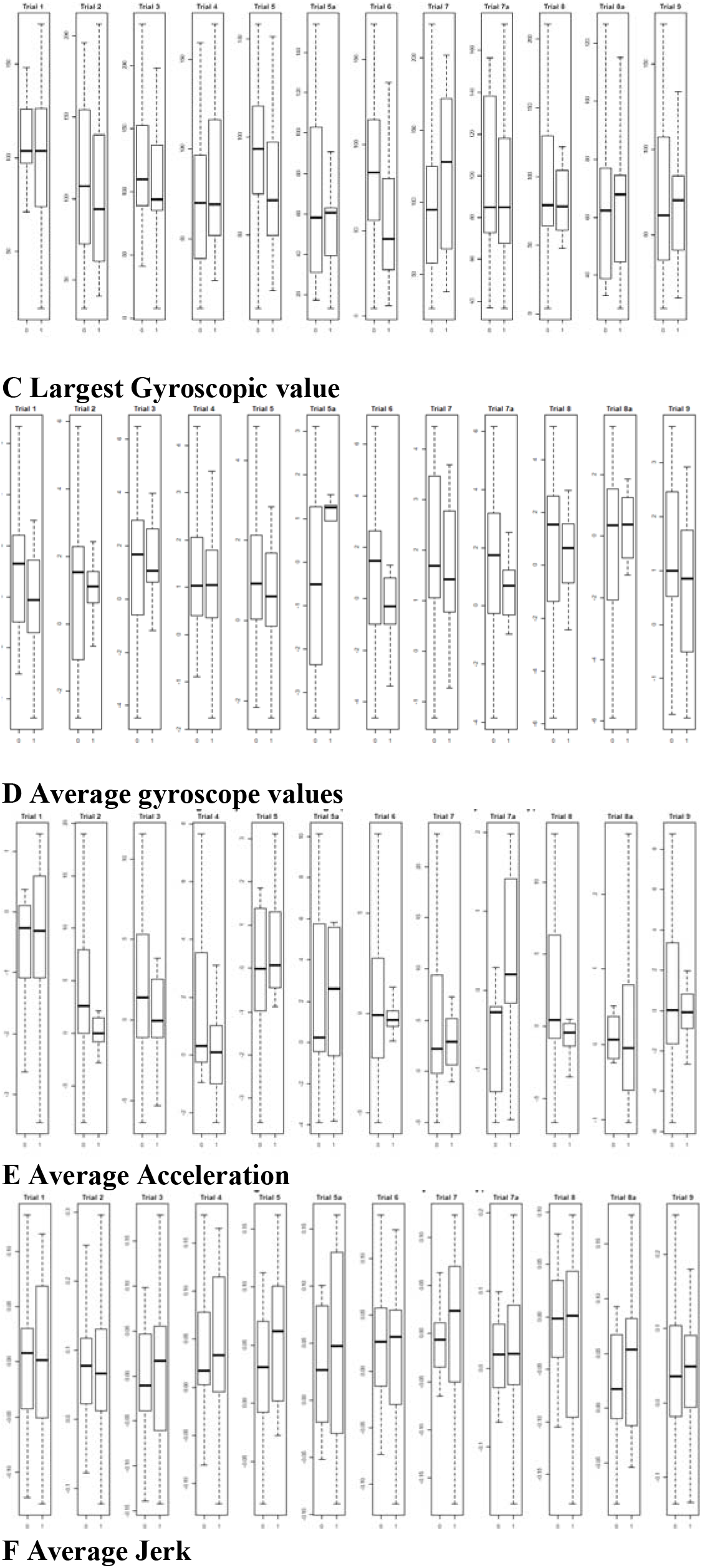
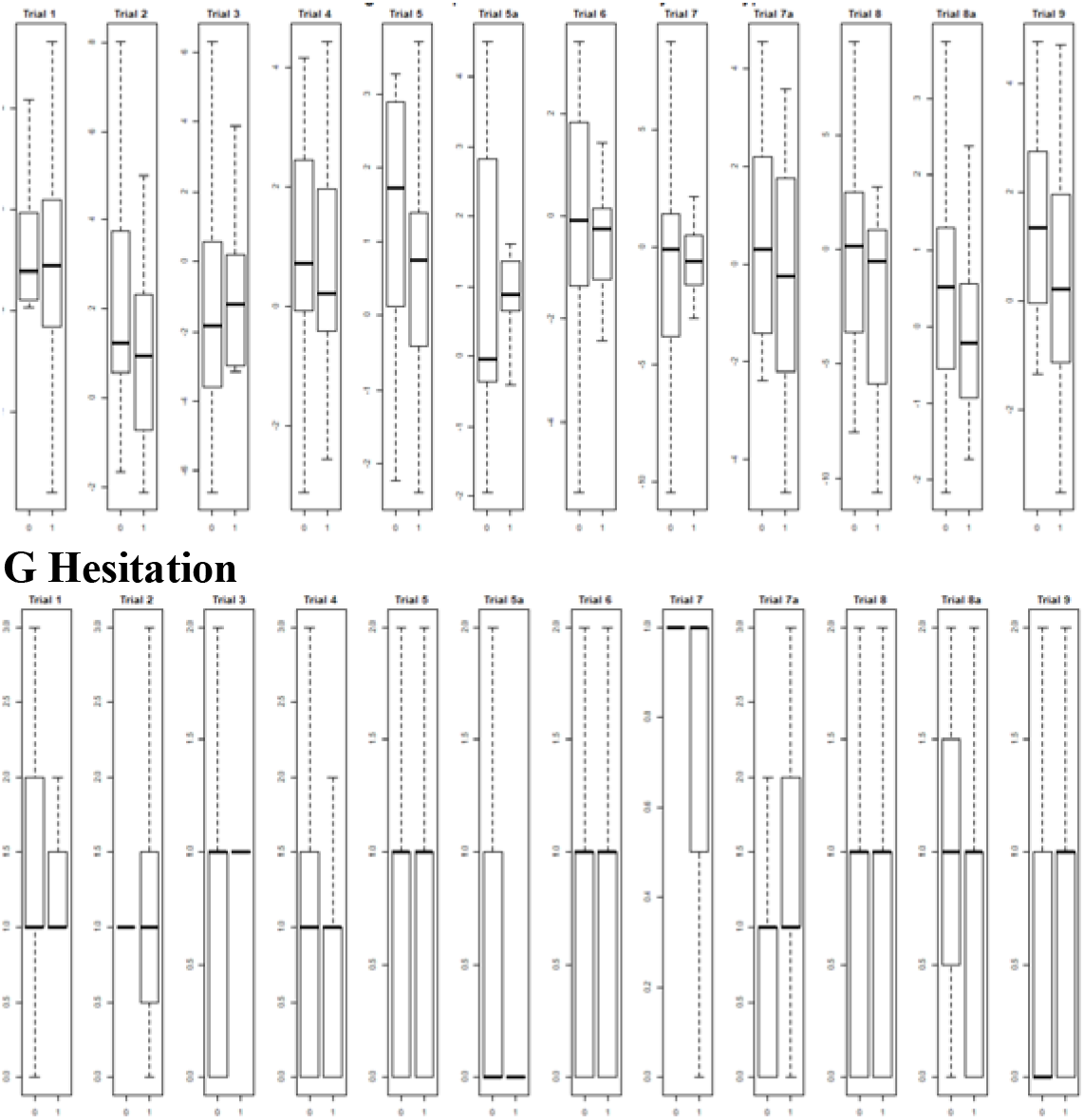
Box plot for each task feature on each trial split by *APOE* genotype and analysed with univariate statistics.

**Table S1.**
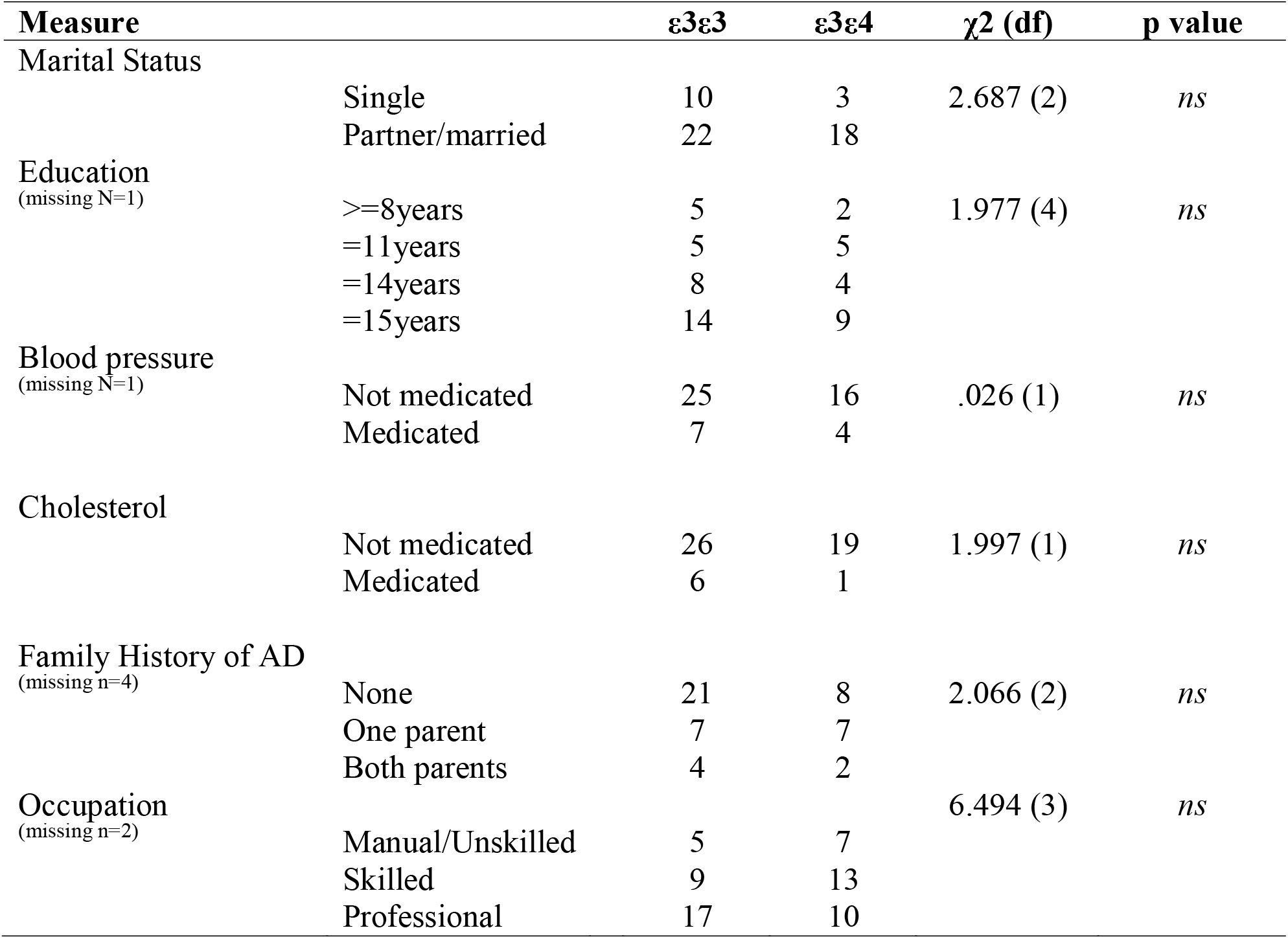
Secondary characteristics of the sample. Secondary characteristics between genetic groups. No difference on any of the above listed characteristics were detected with Persons Chi square confirmed analysis.

| Measure |  | ε3ε3 | ε3ε4 | χ<sup>2</sup> (df) | p value |
| --- | --- | --- | --- | --- | --- |
| Marital Status |  |  |  |  |  |
|  | Single | 10 | 3 | 2.687 (2) | *ns* |
|  | Partner/married | 22 | 18 |  |  |
| Education<br>(missing N=1) |  |  |  |  |  |
|  | >=8years | 5 | 2 | 1.977 (4) | *ns* |
|  | =11years | 5 | 5 |  |  |
|  | =14years | 8 | 4 |  |  |
|  | =15years | 14 | 9 |  |  |
| Blood pressure<br>(missing N=1) |  |  |  |  |  |
|  | Not medicated | 25 | 16 | .026 (1) | *ns* |
|  | Medicated | 7 | 4 |  |  |
| Cholesterol |  |  |  |  |  |
|  | Not medicated | 26 | 19 | 1.997 (1) | *ns* |
|  | Medicated | 6 | 1 |  |  |
| Family History of AD<br>(missing n=4) |  |  |  |  |  |
|  | None | 21 | 8 | 2.066 (2) | *ns* |
|  | One parent | 7 | 7 |  |  |
|  | Both parents | 4 | 2 |  |  |
| Occupation<br>(missing n=2) |  |  |  |  |  |
|  | Manual/Unskilled | 5 | 7 | 6.494 (3) | *ns* |
|  | Skilled | 9 | 13 |  |  |
|  | Professional | 17 | 10 |  |  |

**Table S2.**
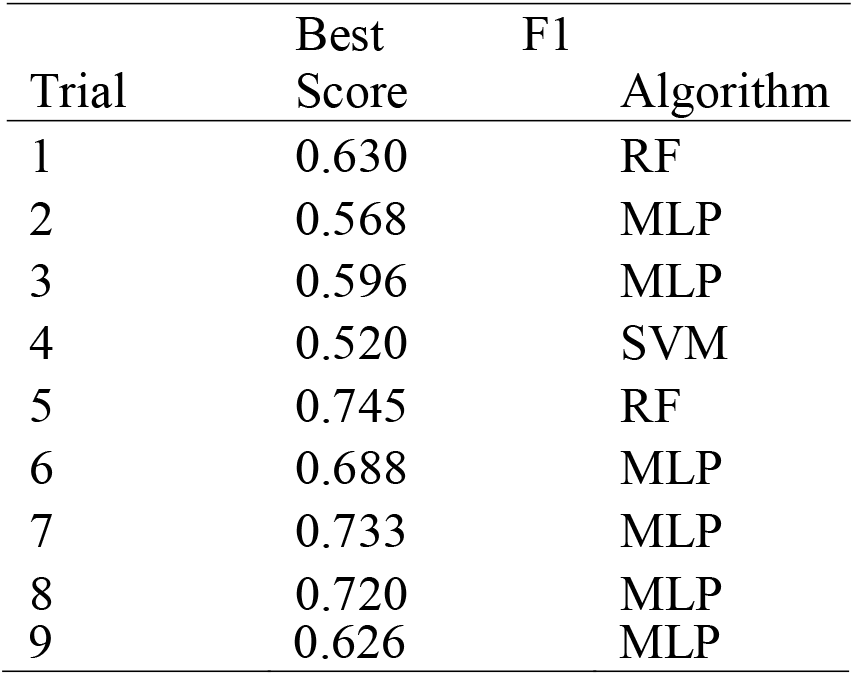
Model performance excluding demographic variables (age, sex and occupation)

**Table S3.** F1 scores reflecting for the best performing prediction models and algorithm on each trial, with ε4ε4 participants included (N=3).

| Trial | Best F1 Score | Algorithm |
| --- | --- | --- |
| 1 | 0.673 | MLP |
| 2 | 0.582 | MLP |
| 3 | 0.587 | MLP |
| 4 | 0.599 | MLP |
| 5 | 0.767 | RF |
| 6 | 0.705 | RF |
| 7 | 0.765 | RF |
| 8 | 0.734 | MLP |
| 9 | 0.703 | MLP |

